# Multicohort development and validation of a machine learning model to predict six-month functional traumatic brain injury outcomes in a large national registry

**DOI:** 10.64898/2026.04.23.26351622

**Authors:** Vikas N. Vattipally, Ritvik R. Jillala, Patrick Kramer, Mazin Elshareif, Shivam Singh, Jacob Jo, Jose I. Suarez, Joseph V. Sakran, Elliott R. Haut, Judy Huang, Chetan Bettegowda, Tej D. Azad

## Abstract

**Background:** Prognostication after moderate-to-severe traumatic brain injury (TBI) rarely captures long-term functional recovery, despite its importance to patients, families, and clinicians. Large trauma registries such as the Trauma Quality Improvement Program (TQIP) dataset contain detailed clinical data but lack systematic follow-up, limiting their ability to study longer-term functional outcomes.

**Methods:** We developed and externally validated a machine learning model to predict favorable six-month functional outcome (GOS “MD”/”GR” or GOSE ≥5) using harmonized data from two randomized clinical trials: CRASH (training) and ROC-TBI (validation). Five candidate classifiers (random forest [RF], linear discriminant analysis, k-nearest neighbors, naïve Bayes, and support vector machine) were trained using seven shared clinical predictors. Models were evaluated using ROC-AUC, calibration metrics, and performance at the Youden optimal threshold and a high-sensitivity secondary threshold. The final model was applied to patients with moderate-to-severe TBI in the national TQIP registry (2017–2022) to estimate population-level recovery patterns.

**Results:** The RF model demonstrated the highest overall performance after recalibration, achieving strong discrimination (AUC internal and external, 0.887 and 0.784), good calibration, and high sensitivity (0.890) and negative predictive value (0.909). Applied to 63,289 patients from TQIP, the model estimated that 45% would achieve favorable six-month outcomes at the Youden optimal threshold and 57% at the high-sensitivity threshold, with predicted recovery aligning with established clinical correlates such as younger age, higher admission GCS, and lower rates of penetrating or brainstem injuries.

**Conclusion:** A machine learning model trained on high-quality trial data can generate clinically plausible estimates of long-term functional recovery when applied at scale to national trauma registries that lack systematic follow-up. This approach enables imputation of functional outcomes in datasets lacking follow-up, supports benchmarking and quality improvement across trauma systems, and provides a foundation for future models incorporating physiologic time-series, imaging, and biomarker data.

## INTRODUCTION

Traumatic brain injury (TBI) remains a leading cause of death and morbidity, with devastating effects on patients and families.^1^ Clinical recovery trajectories vary from full recovery to death or a persistent vegetative state, reflecting both injury heterogeneity and challenges for prognostication.^1–3^ Moreover, traditional predictors such as the Glasgow Coma Scale (GCS) and early imaging findings may provide an initial assessment of injury severity but often demonstrate limited accuracy in forecasting long-term functional outcomes.^4,5^ Increasingly, predictions of long-term functional recovery have become recognized as meaningful for guiding the care of patients with TBI, families, and clinicians.^6^ In this domain, standardized scores such as the Glasgow Outcome Scale (GOS) and GOS-Extended (GOSE) provide operational, patient-centered measures of functional independence at follow-up time points post-injury, corresponding to meaningful recovery trajectories.^7^

Existing data, such as those from the Corticosteroid Randomization after Significant Head injury (CRASH) and Resuscitation Outcomes Consortium-TBI (ROC-TBI) studies, are derived from randomized controlled trials recruiting patients with TBI and have been instrumental for developing longer-term functional prognosis prediction models via GOSE.^8,9^ However, these cohorts include highly-selected patients with limited, specific variables available for analysis, limiting their generalizability to broader trauma cohorts. Large national trauma registries, in contrast, typically provide broad patient representation but lack long-term follow-up data, restricting analyses to in-hospital outcomes or discharge disposition.^10,11^ One such registry, the American College of Surgeons Trauma Quality Improvement Program (TQIP), includes records from hundreds of thousands of patients with TBI treated across hundreds of trauma centers across the United States and Canada, including detailed demographic, comorbidity, injury severity, and treatment data, but no data on longer-term functional outcomes.^12^ Extending functional prognostication models to the TQIP dataset would enable nationwide benchmarking of recovery after TBI, allow assessment of both hospital and system level variation in functional outcomes, and provide more realistic denominators for follow-up and rehabilitation research.

Given the heterogeneity of clinical courses after TBI, traditional prognostication models often exhibit variable calibration and discriminative ability.^13,14^ Machine learning provides a practical solution to this problem, as these models can capture dynamic effects among predictors to learn robust non-linear relationships that can be generalized to broader populations. After training on robust clinical trial data, applying such models to TQIP may yield plausible, probabilistic estimates of six-month functional outcomes, effectively allowing for imputation of follow-up data at scale. Thus, the aim of this study was to develop and validate a machine learning-based model to predict favorable six-month outcomes using data from the CRASH and ROC-TBI trials and apply it to TQIP to estimate patterns of functional recovery among patients in this large registry for use in subsequent retrospective research studies. We hypothesized that model-predicted functional outcomes in TQIP would fall along clinically consistent patterns of patient baseline and clinical variables.

## METHODS

### Study Population

This was a multicenter retrospective model development study leveraging data from multiple datasets. Included patients were those presenting with moderate-to-severe TBI, defined as presenting GCS ≤12, to reflect the TBI populations enrolled in the parent studies and to focus model development on those with greater prognostic uncertainty, thereby enriching the training cohort for meaningful prognostic signal. Ethical approval was provided by the Johns Hopkins University Institutional Review Board (IRB00053752). We first performed model training on data from the CRASH randomized placebo-controlled trial.^15^ This trial recruited 10,008 patients presenting with TBI and GCS ≤14 and randomly assigned them to either receive an infusion of corticosteroids or placebo shortly after injury. The authors found that patients who received corticosteroids experienced greater 2-week mortality and poorer functional outcomes at six months of follow-up.^8^ Next, we validated our model using data from the ROC-TBI randomized controlled trial (N=1,331).^16^ This study sought to determine if administering hypertonic fluids improved six-month functional outcomes after severe TBI (*i.e.*, presenting GCS ≤8) over normal saline infusion, for which the authors observed no beneficial effect. Finally, model predictions were applied to patients with TBI (*i.e.* primary International Classification of Diseases, 10^th^ edition code under the category “S06”) in the TQIP dataset presenting between 2017 and 2022.

Our cohort selection process is detailed in the **Supplemental Figure**. As noted previously, we first selected for patients with moderate-to-severe TBI. Next, we excluded patients who were missing data for shared prognostic indicators of interest that we planned to use in model development. Imputation of these variables was considered but was decided against because performing model training and validation on already-imputed data would risk propagating artificial correlations and the model’s learned relationships. Finally, patients in the CRASH and ROC-TBI cohorts who were missing data for GOS and GOSE at six months, respectively, were excluded.

### Outcome of Interest

We sought to develop our model to predict favorable functional outcome at six months after injury. This was defined as a GOSE score ≥5 in the ROC-TBI cohort, as was operationalized by authors of that parent study.^16^ This corresponds to “moderate disability (MD)” or “good recovery (GR)” at the follow-up time point, as opposed to “severe disability (SD),” “vegetative state (VS),” or death. The CRASH cohort contains GOS data rather than GOSE, so favorable functional outcome was defined as GR or MD as opposed to SD, VS, or death in order to align with the 1-8 scoring of GOSE.

### Shared Predictor Variables

A set of seven patient baseline, injury, treatment, and outcome variables were selected as model predictors for their potential prognostic value for favorable six-month outcome based on clinical experience of the authors, literature review, and shared presence in all three datasets. These variables were age, sex, GCS at presentation, the presence of polytrauma (defined as “major extracranial injury” in CRASH and as an Injury Severity Score ≥16 in ROC-TBI and TQIP),^17,18^ pupil reactivity at presentation (*i.e.,* both, one, or neither), receipt of cranial surgery, and hospital discharge disposition (*i.e.,* home, rehabilitation, other inpatient care, or death) **(Figure 1)**.

**Figure 1.**
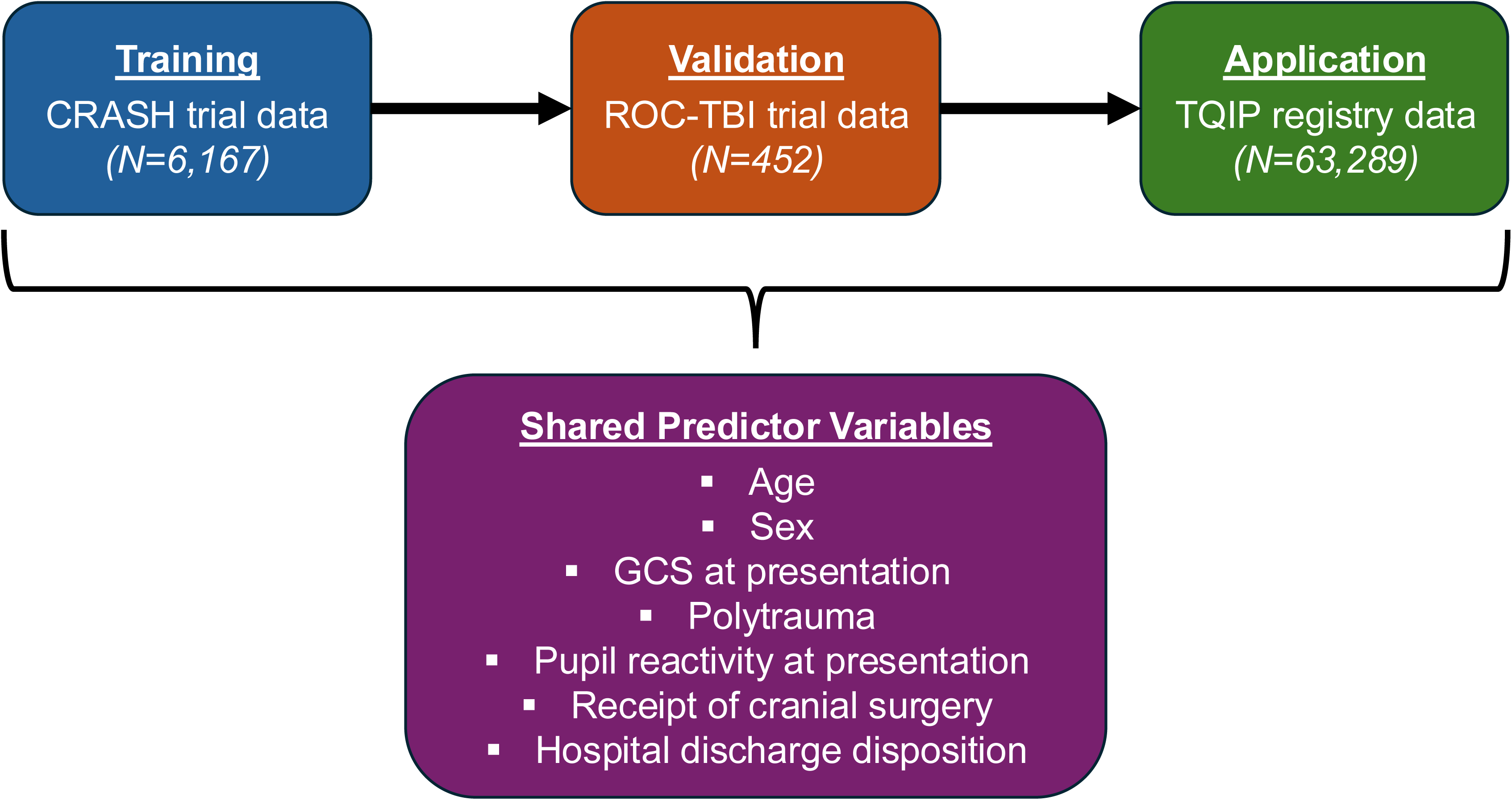
Overview of study cohorts and shared predictor variables. *CRASH – Corticosteroid Randomization after Significant Head injury; ROC – Resuscitation Outcomes Consortium-TBI; TQIP – Trauma Quality Improvement Program*; *GCS – Glasgow Coma Scale*

### Statistical Analysis

Univariable comparisons were performed between the training (CRASH) and validation (ROC) cohorts on the predictor variables and favorable outcome. Standardized mean differences (SMD) were reported, with imbalances defined as an absolute value >0.10. Continuous predictors were centered and scaled using parameters estimated from the training dataset, and the same transformation was later applied to the validation and application datasets. We fitted five candidate machine learning-based classifiers to predict the outcome on the training set based on prior database imputation work: random forest (RF), linear discriminant analysis (LDA), k-nearest neighbors (KNN), naïve Bayes (NB), and radial-basis support vector machine (SVM).^19^ Model tuning used stratified five-fold cross-validation repeated five times with area under the receiver operating characteristic curve (ROC-AUC) as the optimization metric and cross-validated class probabilities retained. To characterize each model, we visualized cross-validated ROC-AUC and classification metrics (sensitivity, specificity, positive predictive value [PPV], negative predictive value [NPV], accuracy) at the Youden optimal threshold.

All five candidate models were evaluated in the ROC-TBI validation cohort using predicted probabilities generated from the CRASH-trained models and CRASH-derived scaling. We computed ROC-AUC, Youden optimal threshold accuracy, Brier score, calibration slope, and calibration intercept for each model and shortlisted the two models with the highest ROC-AUC. For each, we performed logistic recalibration in the validation dataset and recomputed ROC-AUC, Brier score, calibration slope, and calibration intercept. The final model was chosen as the one of these two with the higher post-recalibration ROC-AUC, and a calibration plot was constructed using decile-of-risk binning and Wilson 95% confidence intervals. For the chosen model, we defined the primary operating threshold by Youden’s J statistic on the receiver operating characteristic curve of the validation dataset. At this threshold, we reported the confusion matrix, accuracy, sensitivity, specificity, PPV, and NPV. As a prespecified sensitivity analysis, we also identified a secondary high-sensitivity operating threshold by selecting the lowest probability cut-point that achieved a sensitivity value ≥0.950 for favorable outcome in the validation cohort and summarized model performance at this threshold. Finally, we calculated Shapley Additive Explanation (SHAP) values for each variable and visualized these using beeswarm plots to evaluate relative feature importance.

The validation cohort was then compared to the application TQIP cohort with SMD values. We then applied the CRASH-derived scaling to TQIP variables and applied the chosen model to generate raw probabilities that were then recalibrated and assigned final class by the prespecified threshold. In addition, we applied the secondary high-sensitivity threshold to the recalibrated probabilities in TQIP to obtain a conservative estimate of the proportion of patients predicted to achieve favorable six-month outcomes, effectively “ruling out” patients unlikely to experience favorable recovery. To confirm clinical plausibility, we compared patients in TQIP based on predicted six-month outcome at the original threshold. All analysis was performed in R (version 4.2.2).

## RESULTS

### Training and Validation Cohorts Patient Population

We identified 6,167 patients for inclusion in the CRASH training dataset and 452 in the ROC-TBI validation dataset **(Supplemental Table 1)**. Patients in the validation dataset had more severe neurologic presentation (median GCS, 3 vs. 8; SMD=1.5) and higher rates of cranial surgery (37% vs. 25%; SMD=0.26), but lower rates of home discharge (23% vs. 41%; SMD=1.0) and favorable six-month outcome (34% vs. 53%; SMD=0.38).

### Model Selection

**Supplemental Figure 2** presents internally cross-validated metrics for each of the five candidate models. LDA and RF had the highest ROC-AUC values (0.887 and 0.885, respectively), and RF demonstrated the highest sensitivity (0.884), NPV (0.851), and accuracy (0.817). When these five models were applied to the external cohort, the RF (ROC-AUC=0.784) and LDA (ROC-AUC=0.781) models were selected as the two shortlisted models based on their superior ROC-AUC values **(Table 1**, **Figure 2)**. Logistic recalibration resulted in normalization of the slope and intercept for both models, as expected **(Supplemental Table 2)**. After this recalibration, RF was selected as the top model based on its ROC-AUC of 0.784.

**Figure 2.**
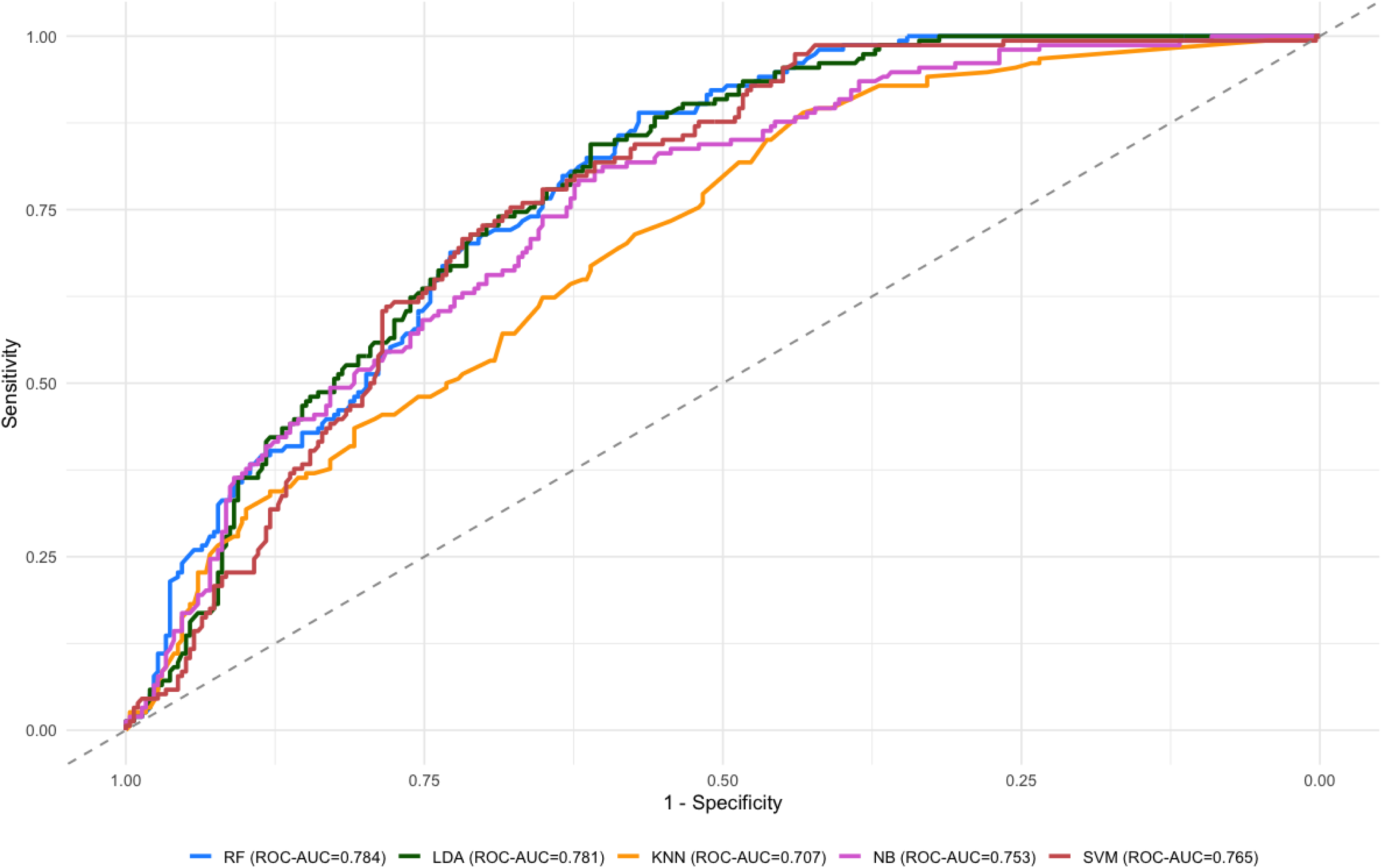
External validation receiver operating characteristic curves for candidate machine learning models. *ROC-AUC – area under the receiver operating characteristic curve; RF – random forest; LDA – linear discriminant analysis; KNN – k-nearest neighbors; NB – naïve Bayes; SVM – support vector machine*

**Table 1.**
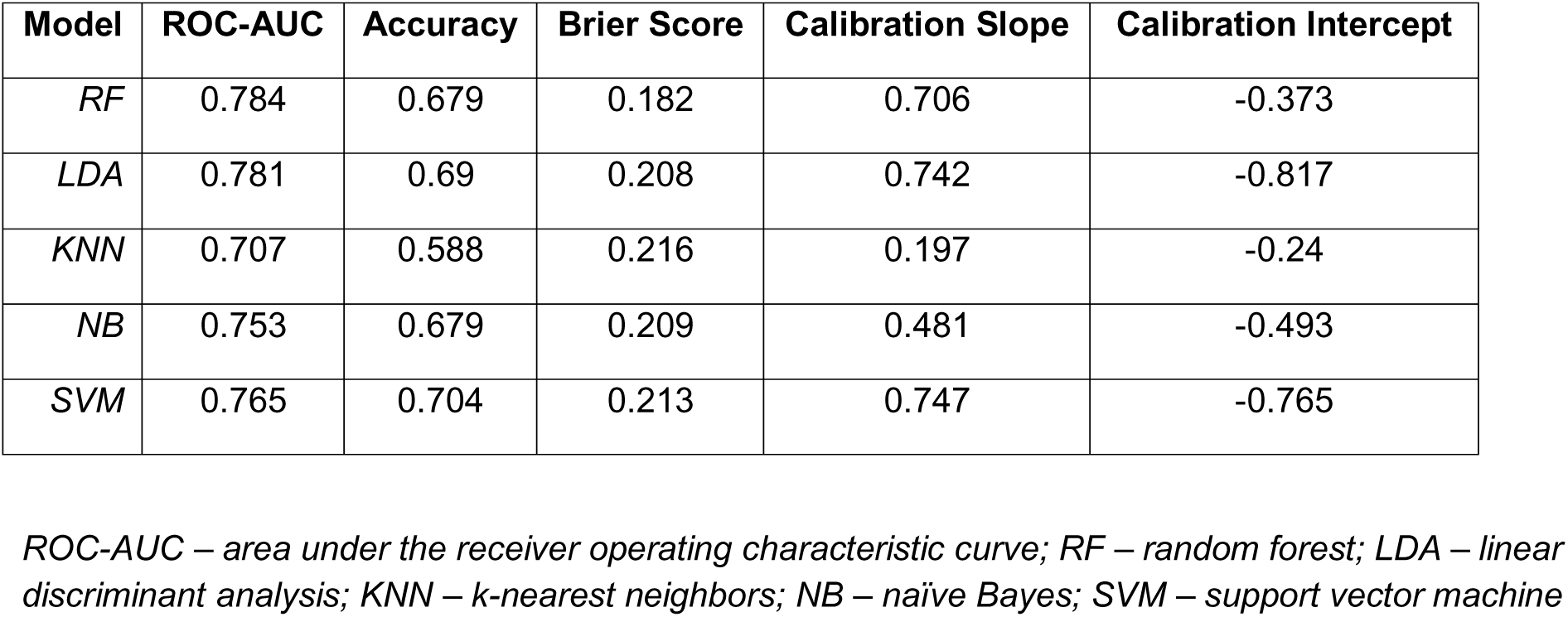
External validation performance of candidate prediction models.

### RF Characterization

The RF calibration plot showed that the predicted and observed risks aligned well for low predicted probabilities, with moderate overestimation at higher predicted probabilities **(Supplemental Figure 3)**. The Youden optimal probability threshold was determined to be 0.312 **(Supplemental Table 4)**. At this threshold, the RF model exhibited strong sensitivity (0.890) and NPV (0.909), as well as moderate accuracy (0.679) and acceptable specificity (0.570) and PPV (0.517). The high-sensitivity secondary analysis revealed a threshold of 0.229, with a resulting sensitivity of 0.955 and accuracy of 0.620. From the SHAP beeswarm plot **(Figure 3)**, the most important features towards predicting six-month favorable outcomes were determined to be hospital discharge disposition and GCS at presentation.

**Figure 3.**
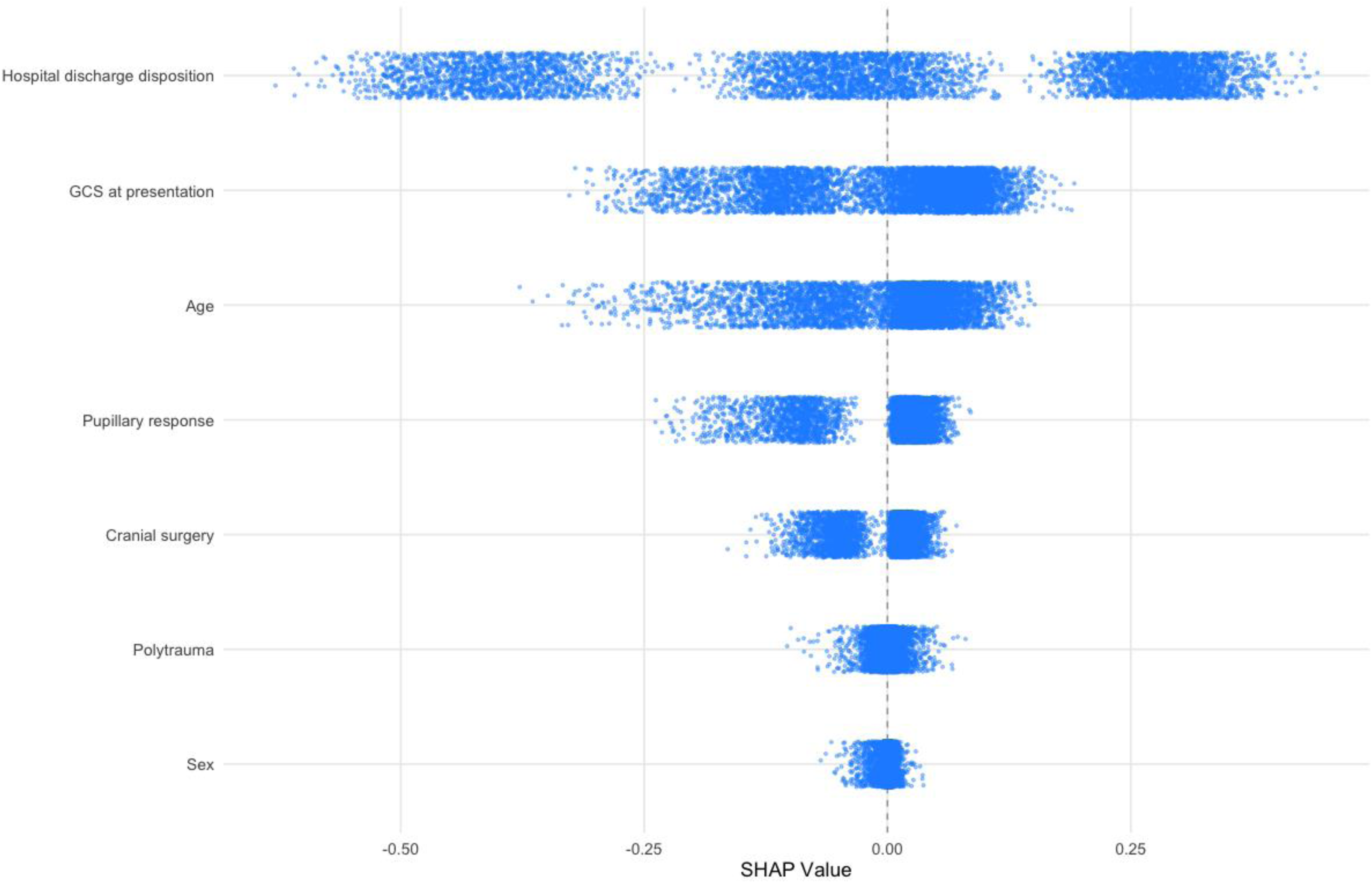
Shapley Additive Explanation values ranked by feature importance for the final random forest model. *GCS – Glasgow Coma Scale*

### Application Cohort Patient Population

A total of 63,289 patients were identified for model application from TQIP **(Table 2)**. Compared to patients in the validation cohort, those from TQIP were older (median age, 49 years vs. 32 years; SMD=-0.55), less neurologically impaired (median GCS, 4 vs. 3; SMD=-0.64), and had lower rates of cranial surgery (20% vs. 37%; SMD=0.37) but higher rates of inpatient mortality (29% vs 21%; SMD=0.50).

**Table 2.**
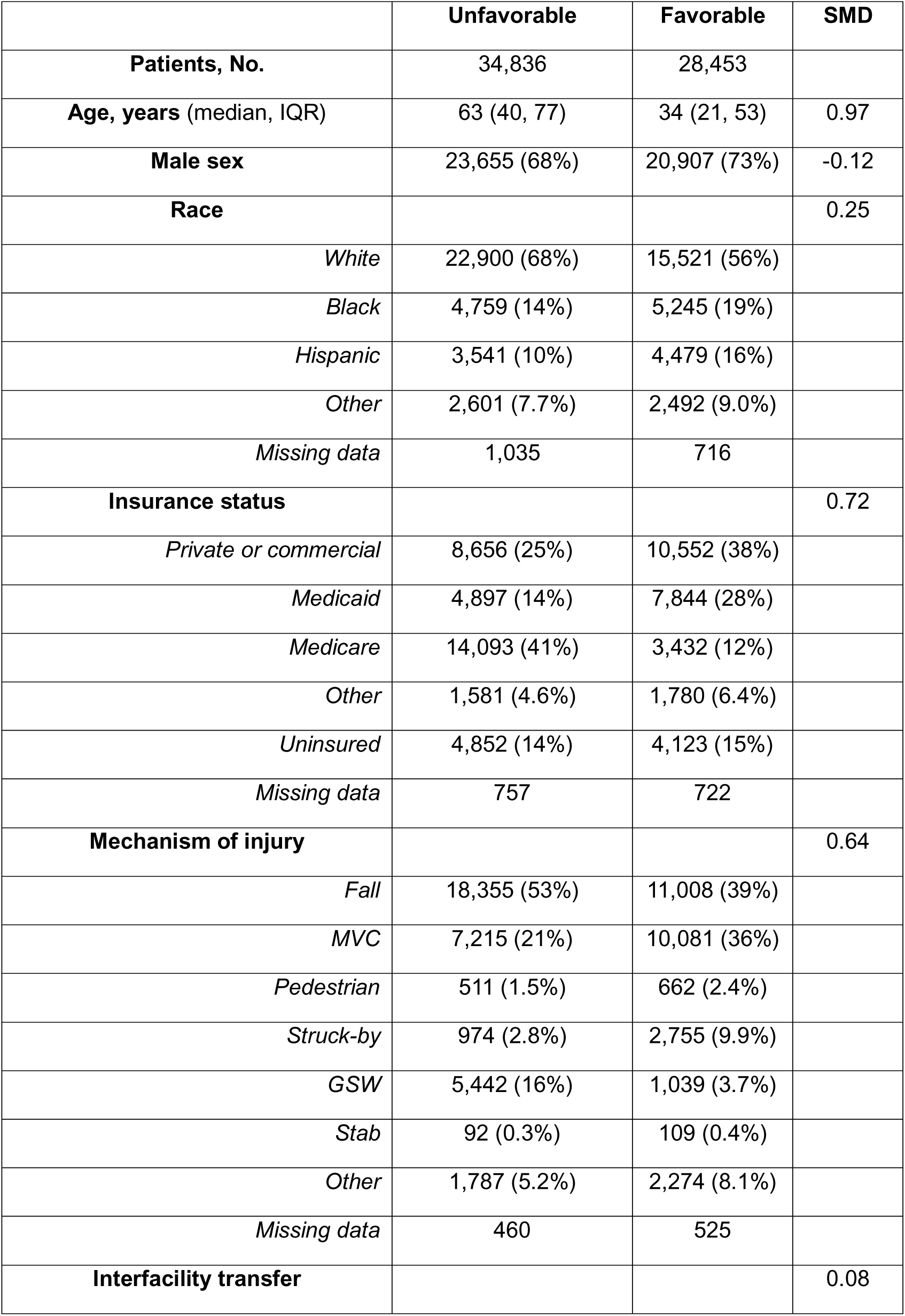
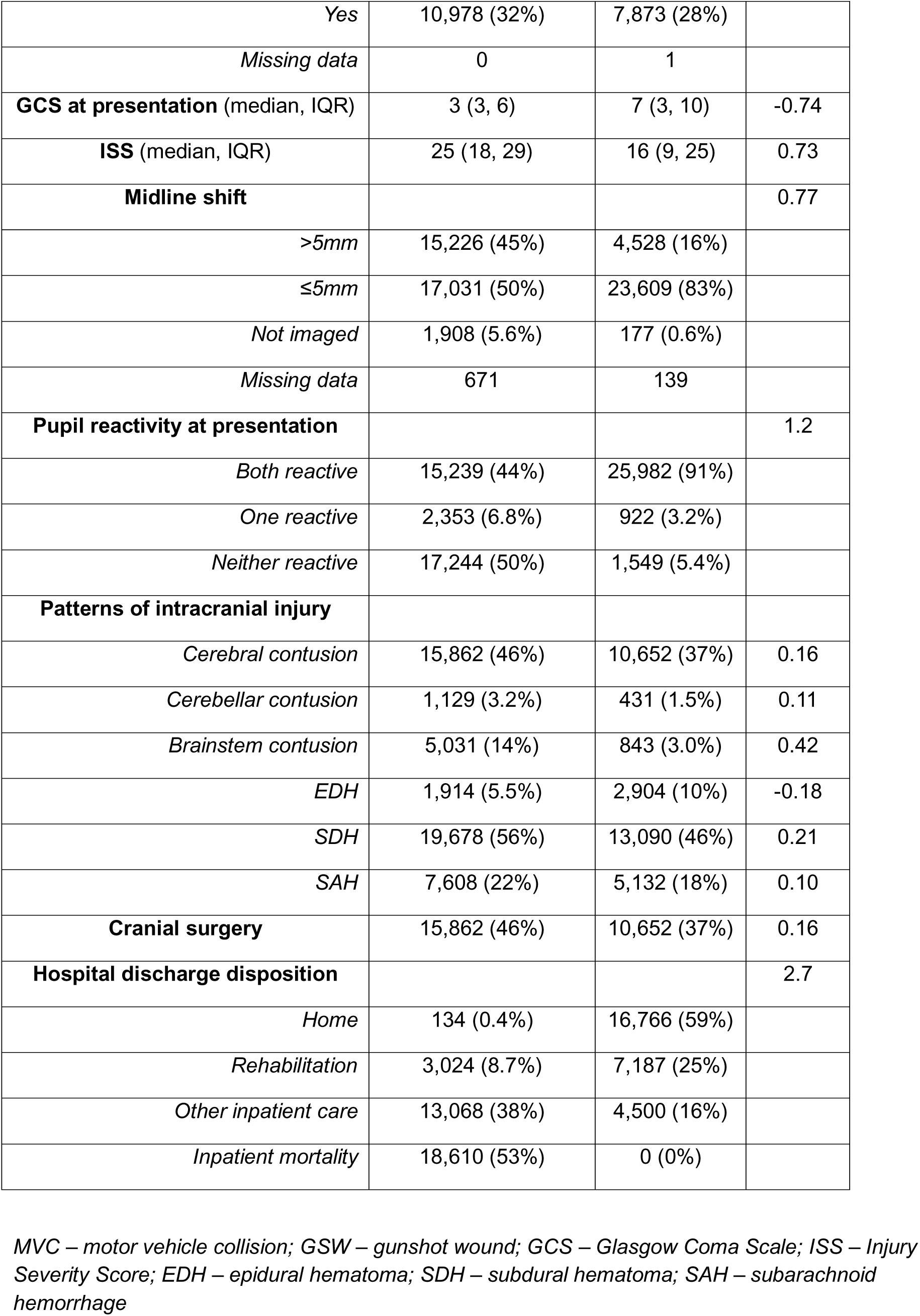
Characteristics of patients with TBI in TQIP database, stratified by predicted favorable six-month outcome.

Applying the RF model and recalibrated predictions to the TQIP dataset revealed 28,453 patients (45%) who achieved a favorable six-month outcome at the Youden optimal threshold **(Supplemental Table 4)**. As expected, these patients were younger (median age, 34 years vs. 63 years) with lower rates of high-impact mechanisms such as gunshot wounds (3.7% vs. 16%; SMD=0.64) and patterns of intracranial injury such as brainstem contusions (3.0% vs. 14%; SMD=0.42). At the high-sensitivity threshold, the proportion of patients with predicted favorable outcomes increased to 57% (N=35,821).

## DISCUSSION

Using data from two multicenter, randomized controlled trial cohorts, we derived and validated a machine learning model to robustly predict six-month favorable recovery among patients with moderate-to-severe TBI. Among five candidate algorithms, RF demonstrated the best overall performance after recalibration, with good agreement at low-to-moderate predicted probabilities and modest overestimation at higher risk levels. At its optimal probability threshold, RF yielded high sensitivity (0.890), strong NPV (0.909), and moderate accuracy (0.679). Finally, when applied to the national TQIP registry, our model estimated a 45% rate of favorable outcomes, aligning with expected clinical correlates including younger age, higher admission GCS, and lower rates of firearm and brainstem injury.^10,20–22^ Moreover, we evaluated a high-sensitivity (≥0.950) operating point to reflect a more conservative rule-out strategy for favorable recovery. When this threshold was applied to TQIP, the proportion of patients predicted to regain favorable function increased to 57%, providing an upper-bound estimate of potential recovery. These longer-term functional recovery rates align broadly with those previously reported in a TRACK-TBI cohort.^23^

The final RF model demonstrated flexibility, enabling it to capture nonlinear associations among key predictors such as admission GCS, age, pupillary response, and need for cranial surgery. It demonstrated strong discrimination across both the derivation and validation datasets (AUC internal and external, 0.887 and 0.784), supporting internal validity and robust generalizability. Notably, performance remained stable despite substantial clinical and distributional differences between cohorts, including injury severity profiles, intervention rates, and outcome prevalence, underscoring the model’s robustness to dataset heterogeneity. After recalibration, the calibration slope approximated 1 and intercept approached 0, indicating well-scaled predictions with minimal systematic bias. Calibration curves showed excellent alignment between predicted and observed risks across most of the probability range, with mild overestimation at higher predicted risks likely reflecting sparse observations.

Clinicians are generally limited in their ability to perform prognostication for patients with moderate-to-severe TBI beyond short-term mortality outcomes, often struggling to address the consequential question of who ultimately regains function.^24,25^ Machine learning-based prediction models for longer-term functional scores such as GOSE directly target this broader horizon, capturing meaningful patient-centered outcomes that mortality alone cannot reflect such as independence, community reintegration, and return to work.^26,27^ The application of our model to over 60,000 patients with TBI in the TQIP registry directly connects high-quality but narrow clinical trials with large, representative trauma databases. This approach aligns with recommendations to incorporate long-term outcomes into trauma registries for benchmarking care quality and improving clinical outcomes research.^28^

This work demonstrates the feasibility of model-based imputation of long-term functional outcomes within large trauma registries that lack systematic follow-up. Integration of this model could enable national benchmarking of TBI recovery, support the development of risk-adjusted performance metrics within TQIP, and broaden the scope of quality improvement efforts centered on functional, not merely survival, outcomes. Future work should incorporate time-series physiologic data, imaging-derived features, and emerging blood-based biomarkers to further refine model estimates. Prospective validation of model-imputed functional outcomes against true six-month follow-up data will be essential.

### Limitations

This study has several limitations. First, heterogeneity in variable definitions across datasets may introduce misalignment, specifically polytrauma being defined as “major extracranial injury” in CRASH and approximated using ISS ≥16 in ROC-TBI and TQIP, and the harmonization of GOS with GOSE reducing the granularity of functional outcome categories. Second, both CRASH and ROC-TBI enrolled trial-eligible patients, introducing selection bias that may underrepresent mild, non-operative, or highly comorbid TBI populations. Third, only seven shared predictors were available across cohorts, limiting the incorporation of imaging features, physiologic time-series data, or biomarker profiles. Additionally, cross-cohort differences in injury patterns, case mix, and care processes between CRASH, ROC, and TQIP may constrain external transportability, and temporal drift, given that the CRASH and ROC trials were conducted more than a decade prior to modern TQIP. Finally, although the random forest achieved the strongest performance, its ensemble structure limits direct interpretability relative to simpler regression models.^29^

## CONCLUSION

In this multi-cohort study, we developed, validated, and applied a machine learning model capable of estimating six-month functional recovery after moderate-to-severe TBI using harmonized data from randomized trials and a national trauma registry. The final RF model demonstrated strong discrimination, good calibration after recalibration, and high sensitivity and NPV. When extended to TQIP, the model generated clinically plausible, population-level estimates of functional recovery, illustrating how trial-derived prognostic tools can be leveraged to enhance benchmarking and support quality improvement in real-world trauma systems. By enabling large-scale imputation of functional outcomes for patients with TBI, this approach helps bridge the gap between high-quality clinical trials and large-scale registries lacking follow-up, offering a foundation for future work integrating richer physiologic, imaging, and biomarker data.

## Data Availability

All data produced in the present study are available upon reasonable request to the authors

## Previous Presentations

none

## SUPPLEMENTAL FIGURE LEGENDS

**Supplemental Figure 1.** Multi-cohort flow diagram. CRASH – Corticosteroid Randomization after Significant Head injury; ROC – Resuscitation Outcomes Consortium-TBI; TQIP – Trauma Quality Improvement Program; GCS – Glasgow Coma Scale

**Supplemental Figure 2.** Internal cross-validated performance of candidate machine learning models. ROC-AUC – area under the receiver operating characteristic curve; PPV – positive predictive value; NPV – negative predictive value; KNN – k-nearest neighbors; LDA – linear discriminant analysis; NB – naïve Bayes; RF – random forest; SVM – support vector machine

**Supplemental Figure 3.** Calibration curve for recalibrated random forest model with decile-based observed outcomes and Wilson 95% confidence intervals.

