## Supplementary figures and images for "Multicohort development and validation of a machine learning model to predict six-month functional traumatic brain injury outcomes in a large national registry"

### Supplemental Figures

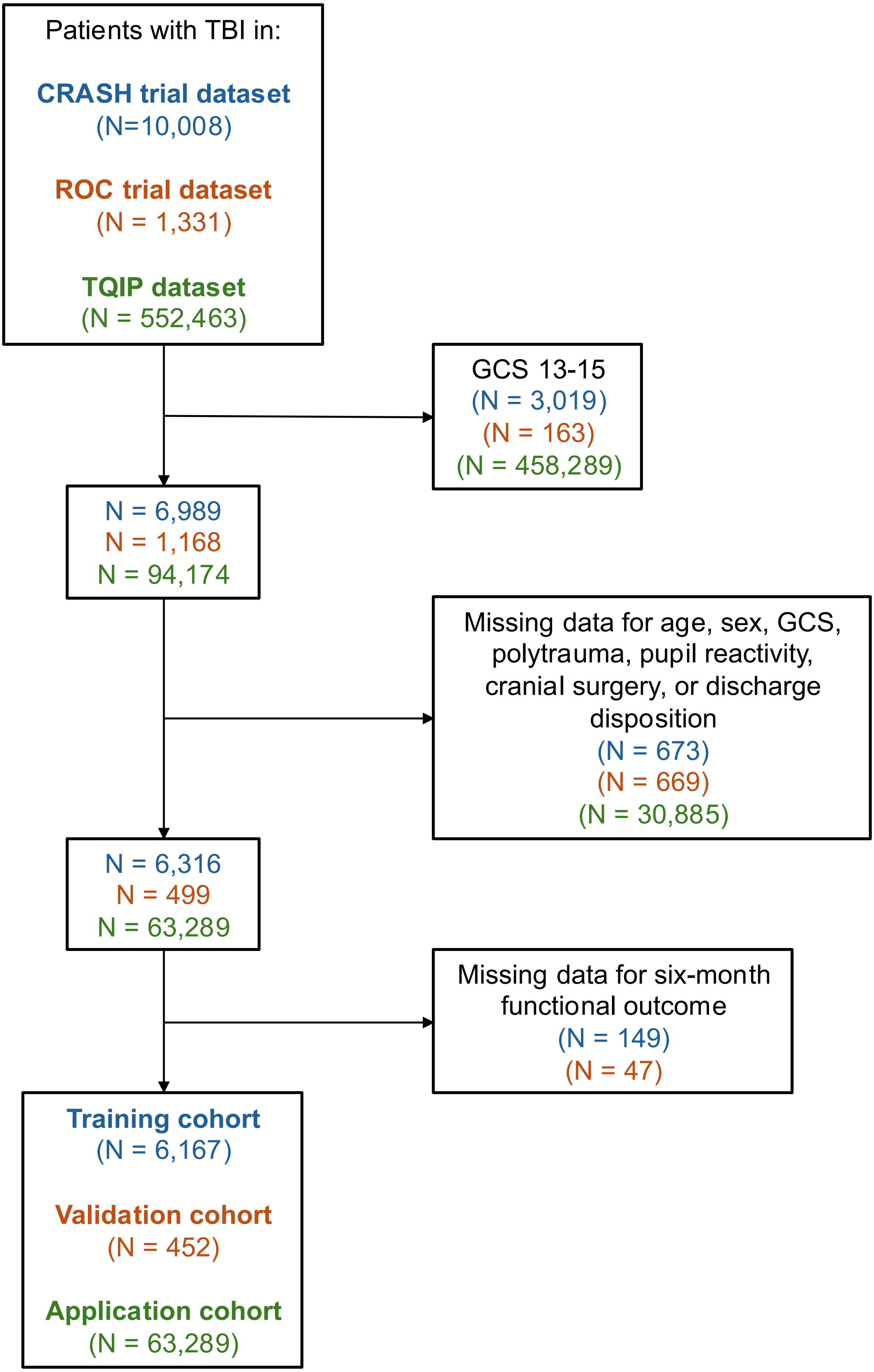

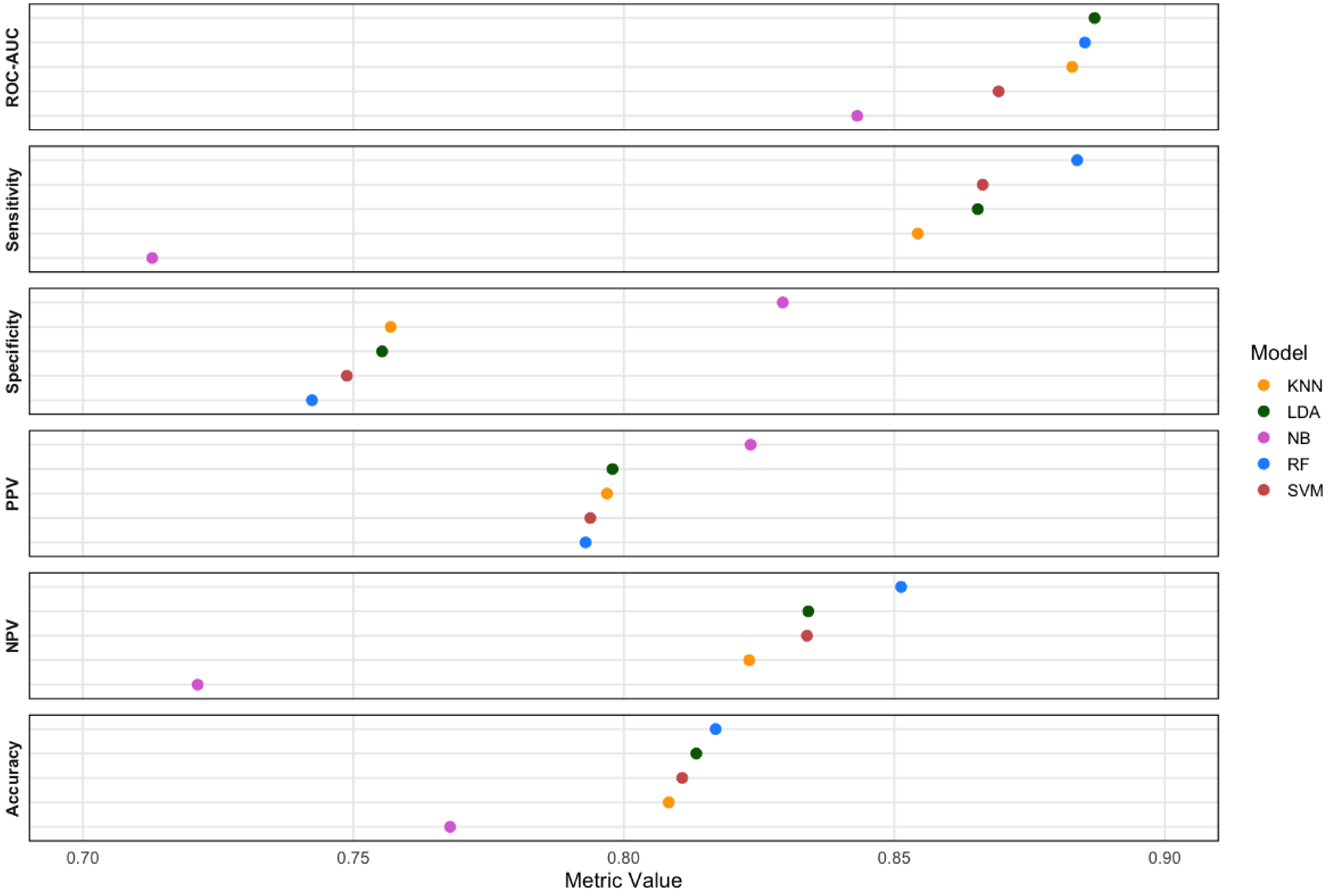

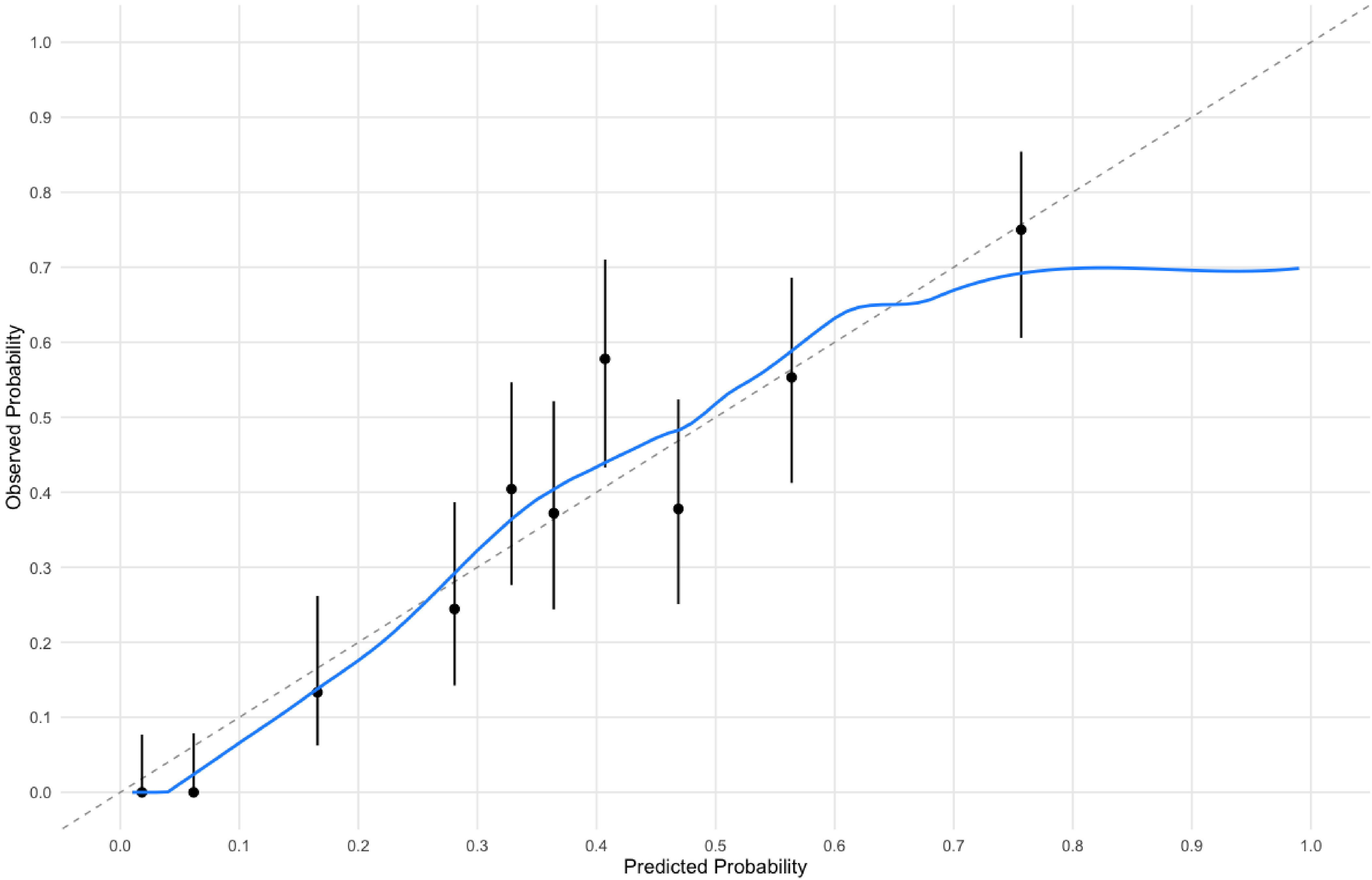
