## Supplemental Tables for "Multicohort development and validation of a machine learning model to predict six-month functional traumatic brain injury outcomes in a large national registry"

**Supplemental Table 1.** Characteristics of patients with TBI compared across training and validation data sources.

|  | **CRASH** (training) | **ROC** (validation) | **SMD** |
| --- | --- | --- | --- |
| **Patients, No.** | 6,167 | 452 |  |
| **Age, years** (median, IQR) | 32 (23, 46) | 32 (24, 48) | -0.04 |
| **Male sex** | 5,101 (83%) | 339 (75%) | 0.19 |
| **GCS at presentation** (median, IQR) | 8 (6, 11) | 3 (3, 5) | 1.5 |
| **Polytrauma** | 1,539 (25%) | 428 (95%) | 2.0 |
| **Pupil reactivity at presentation** |  |  | 0.49 |
| *Both reactive* | 4,900 (79%) | 276 (61%) |  |
| *One reactive* | 515 (8.4%) | 31 (6.9%) |  |
| *Neither reactive* | 752 (12%) | 145 (32%) |  |
| **Cranial surgery** | 1,526 (25%) | 165 (37%) | 0.26 |
| **Hospital discharge disposition** |  |  | 1.0 |
| *Home* | 2,548 (41%) | 106 (23%) |  |
| *Rehabilitation* | 131 (2.1%) | 168 (37%) |  |
| *Other inpatient care* | 1,929 (31%) | 84 (19%) |  |
| *Inpatient mortality* | 1,559 (25%) | 94 (21%) |  |
| **Favorable six-month outcome** | 3,253 (53%) | 154 (34%) | 0.38 |

*GCS – Glasgow Coma Scale*

**Supplemental Table 2.** Calibration metrics of top two candidate prediction models before and after logistic recalibration in validation cohort.

| **Model** | **Phase** | **ROC-AUC** | **Brier Score** | **Calibration Slope** | **Calibration Intercept** |
| --- | --- | --- | --- | --- | --- |
| *RF* | Pre-recalibration | 0.784 | 0.182 | 0.706 | -0.373 |
|  | Post-recalibration | 0.784 | 0.178 | 1.000 | 0 |
| *LDA* | Pre-recalibration | 0.781 | 0.208 | 0.742 | -0.817 |
|  | Post-recalibration | 0.781 | 0.177 | 1.000 | 0 |

*ROC-AUC – area under the receiver operating characteristic curve; RF – random forest; LDA – linear discriminant analysis*

**Supplemental Table 3.** Confusion matrix and performance metrics for top candidate model (random forest) applied to validation cohort at the Youden optimal threshold and minimum threshold that allows for a sensitivity greater than 0.95.

| **Youden Optimal Threshold** | | | | **≥0.95 Sensitivity Threshold** | | |
| --- | --- | --- | --- | --- | --- | --- |
|  | **Observed** | | |  | **Observed** | |
| **Predicted** | *Unfavorable* | | *Favorable* | **Predicted** | *Unfavorable* | *Favorable* |
| *Unfavorable* | 170 | | 17 | *Unfavorable* | 133 | 7 |
| *Favorable* | 128 | | 137 | *Favorable* | 165 | 147 |
| **Performance Metrics** | | | | **Performance Metrics** | | |
| *Threshold* | | 0.312 | | *Threshold* | 0.229 | |
| *Accuracy* | | 0.679 | | *Accuracy* | 0.620 | |
| *Sensitivity* | | 0.890 | | *Sensitivity* | 0.955 | |
| *Specificity* | | 0.570 | | *Specificity* | 0.446 | |
| *PPV* | | 0.517 | | *PPV* | 0.471 | |
| *NPV* | | 0.909 | | *NPV* | 0.950 | |

*PPV – positive predictive value; NPV – negative predictive value*

**Supplemental Table 4.** Characteristics of patients with TBI compared across validation and application data sources.

|  | **ROC** (training) | **TQIP** (application) | **SMD** |
| --- | --- | --- | --- |
| **Patients, No.** | 452 | 63,289 |  |
| **Age, years** (median, IQR) | 32 (24, 48) | 49 (27, 69) | -0.55 |
| **Male sex** | 339 (75%) | 44,562 (70%) | 0.10 |
| **GCS at presentation** (median, IQR) | 3 (3, 5) | 4 (3, 9) | -0.64 |
| **Polytrauma** | 428 (95%) | 45,058 (71%) | 0.66 |
| **Pupil reactivity at presentation** |  |  | 0.10 |
| *Both reactive* | 276 (61%) | 41,221 (65%) |  |
| *One reactive* | 31 (6.9%) | 3,275 (5.2%) |  |
| *Neither reactive* | 145 (32%) | 18,793 (30%) |  |
| **Cranial surgery** | 165 (37%) | 12,740 (20%) | 0.37 |
| **Hospital discharge disposition** |  |  | 0.50 |
| *Home* | 106 (23%) | 16,900 (27%) |  |
| *Rehabilitation* | 168 (37%) | 10,211 (16%) |  |
| *Other inpatient care* | 84 (19%) | 17,568 (28%) |  |
| *Inpatient mortality* | 94 (21%) | 18,610 (29%) |  |

*GCS – Glasgow Coma Scale*
