## Supplementary material for "Multicohort development and validation of a machine learning model to predict six-month functional traumatic brain injury outcomes in a large national registry": Tables

**Table 1.** External validation performance of candidate prediction models.

| **Model** | **ROC-AUC** | **Accuracy** | **Brier Score** | **Calibration Slope** | **Calibration Intercept** |
| --- | --- | --- | --- | --- | --- |
| *RF* | 0.784 | 0.679 | 0.182 | 0.706 | -0.373 |
| *LDA* | 0.781 | 0.69 | 0.208 | 0.742 | -0.817 |
| *KNN* | 0.707 | 0.588 | 0.216 | 0.197 | -0.24 |
| *NB* | 0.753 | 0.679 | 0.209 | 0.481 | -0.493 |
| *SVM* | 0.765 | 0.704 | 0.213 | 0.747 | -0.765 |

*ROC-AUC – area under the receiver operating characteristic curve; RF – random forest; LDA – linear discriminant analysis; KNN – k-nearest neighbors; NB – naïve Bayes; SVM – support vector machine*

**Table 2.** Characteristics of patients with TBI in TQIP database, stratified by predicted favorable six-month outcome.

|  | **Unfavorable** | **Favorable** | **SMD** |
| --- | --- | --- | --- |
| **Patients, No.** | 34,836 | 28,453 |  |
| **Age, years** (median, IQR) | 63 (40, 77) | 34 (21, 53) | 0.97 |
| **Male sex** | 23,655 (68%) | 20,907 (73%) | -0.12 |
| **Race** |  |  | 0.25 |
| *White* | 22,900 (68%) | 15,521 (56%) |  |
| *Black* | 4,759 (14%) | 5,245 (19%) |  |
| *Hispanic* | 3,541 (10%) | 4,479 (16%) |  |
| *Other* | 2,601 (7.7%) | 2,492 (9.0%) |  |
| *Missing data* | 1,035 | 716 |  |
| **Insurance status** |  |  | 0.72 |
| *Private or commercial* | 8,656 (25%) | 10,552 (38%) |  |
| *Medicaid* | 4,897 (14%) | 7,844 (28%) |  |
| *Medicare* | 14,093 (41%) | 3,432 (12%) |  |
| *Other* | 1,581 (4.6%) | 1,780 (6.4%) |  |
| *Uninsured* | 4,852 (14%) | 4,123 (15%) |  |
| *Missing data* | 757 | 722 |  |
| **Mechanism of injury** |  |  | 0.64 |
| *Fall* | 18,355 (53%) | 11,008 (39%) |  |
| *MVC* | 7,215 (21%) | 10,081 (36%) |  |
| *Pedestrian* | 511 (1.5%) | 662 (2.4%) |  |
| *Struck-by* | 974 (2.8%) | 2,755 (9.9%) |  |
| *GSW* | 5,442 (16%) | 1,039 (3.7%) |  |
| *Stab* | 92 (0.3%) | 109 (0.4%) |  |
| *Other* | 1,787 (5.2%) | 2,274 (8.1%) |  |
| *Missing data* | 460 | 525 |  |
| **Interfacility transfer** |  |  | 0.08 |
| *Yes* | 10,978 (32%) | 7,873 (28%) |  |
| *Missing data* | 0 | 1 |  |
| **GCS at presentation** (median, IQR) | 3 (3, 6) | 7 (3, 10) | -0.74 |
| **ISS** (median, IQR) | 25 (18, 29) | 16 (9, 25) | 0.73 |
| **Midline shift** |  |  | 0.77 |
| *>5mm* | 15,226 (45%) | 4,528 (16%) |  |
| *≤5mm* | 17,031 (50%) | 23,609 (83%) |  |
| *Not imaged* | 1,908 (5.6%) | 177 (0.6%) |  |
| *Missing data* | 671 | 139 |  |
| **Pupil reactivity at presentation** |  |  | 1.2 |
| *Both reactive* | 15,239 (44%) | 25,982 (91%) |  |
| *One reactive* | 2,353 (6.8%) | 922 (3.2%) |  |
| *Neither reactive* | 17,244 (50%) | 1,549 (5.4%) |  |
| **Patterns of intracranial injury** |  |  |  |
| *Cerebral contusion* | 15,862 (46%) | 10,652 (37%) | 0.16 |
| *Cerebellar contusion* | 1,129 (3.2%) | 431 (1.5%) | 0.11 |
| *Brainstem contusion* | 5,031 (14%) | 843 (3.0%) | 0.42 |
| *EDH* | 1,914 (5.5%) | 2,904 (10%) | -0.18 |
| *SDH* | 19,678 (56%) | 13,090 (46%) | 0.21 |
| *SAH* | 7,608 (22%) | 5,132 (18%) | 0.10 |
| **Cranial surgery** | 15,862 (46%) | 10,652 (37%) | 0.16 |
| **Hospital discharge disposition** |  |  | 2.7 |
| *Home* | 134 (0.4%) | 16,766 (59%) |  |
| *Rehabilitation* | 3,024 (8.7%) | 7,187 (25%) |  |
| *Other inpatient care* | 13,068 (38%) | 4,500 (16%) |  |
| *Inpatient mortality* | 18,610 (53%) | 0 (0%) |  |

*MVC – motor vehicle collision; GSW – gunshot wound; GCS – Glasgow Coma Scale; ISS – Injury Severity Score; EDH – epidural hematoma; SDH – subdural hematoma; SAH – subarachnoid hemorrhage*
